# Assessment of effectiveness of ‘Nilee Vyema brochure’ on knowledge of breastfeeding, kangaroo mother care and hygiene practice among mothers in central Tanzania. A quasi experiment study protocol

**DOI:** 10.1101/2024.09.24.24314283

**Authors:** Shimwe Amos Ngeze, Julius Ntwenya, Richard Mongi

## Abstract

**Background:** Prematurity is a challenge affecting child health worldwide and mothers who are the sole care providers of preterm infants post discharge must possess the necessary knowledge to be able to properly care for their preterm infant at home.

**Objective:** The main objective of this study will be to assess the effectiveness of *NILEE VYEMA* brochure on knowledge of breastfeeding, kangaroo care and hygiene among mothers with preterm infants in central Tanzania. A sample of 112 mothers with preterm infants will be recruited from two randomly selected regional hospitals in central Tanzania in 2024. A quasi pre-test post-test experimental design will be used and the *Nilee Vyema* brochure will be adapted from several literatures. Both the control group and intervention group will be given the pretest questionnaire to assess baseline knowledge. The control group will receive routine education lecture based on breastfeeding, kangaroo mother care and hygiene topics while the intervention group will receive both routine education in conjunction with the *Nilee Vyema* brochure. Interventional sessions will include lectures on breastfeeding, kangaroo mother care and hygiene to influence the knowledge and skills of these mothers in caring for their preterm. After the intervention, both groups will be given the pretest questionnaire which will assess end-line knowledge.

**Analysis:** Data will be analyzed using SPSS (version 20) for descriptive statistics to describe the characteristics of the study samples and inferential statistics to test the effectiveness of the brochure before and after intervention. Furthermore, chi-square test will be used to describe the relationship between categorical variables at p < 0.05.

## Introduction

Prematurity is a challenge affecting child health worldwide. Globally, 14.84 million babies were preterm births (1). Over sixty percent of these premature deaths occur in Sub-Saharan Africa and Southeast Asia (2). Some infants are born, at term, healthy and are prone to minimum health risks while other infants are born before term and are known as pre term infants. Preterm infants are at high risk to disease and poor prognosis. Preterm birth occurs when an infant is born before 37 completed weeks of gestation, a condition that places an infant at significant risk of developing physical and neurodevelopmental disabilities and death (3).

Mothers of premature infants tend to have extended lengths of stay in the hospital setting. These mothers are not discharged until the neonate is stable. However; despite the relative improvement of the neonatal health status and the relative stabilization of their conditions prior to discharge, these babies may experience unstable health conditions at home and experience post-discharge complications (4).

The most important mental health needs for mothers include proximity, assurance, and information (5). Parents are always looking for information about their baby’s progress in neonatal care to ease feelings of shock, disbelief, and uncertainty (6). Educating mothers on their infant care during the hospital stay is linked to better infants’ health and mother-infant interaction outcomes (7). Empowering parents with knowledge to participate in the delivery of care is one of the effective methods of preventing harm associated with post-discharge consequences.

During the hospital stay mothers are verbally taught the basics of infant care such as breastfeeding, changing diapers, measuring body temperature, and bathing which in turn they can incorporate these new skills at home (7). The required necessary information about preterm infants are not always met by the NICU staff due to staff shortages and other high demands placed on nurses and physicians (8). This suggest that, the information given to patients verbally should be supplemented and reinforced with written and illustrated materials to increase the effectiveness of health education. Patients appreciate the usefulness of written materials (9).

Educating a mother of a premature baby and providing a structured written educational information can enhance better understanding and practice (10). A study conducted in Palestine emphasizes the necessity of thoughtful exchange of health information between team members and mothers and establishing pre- and post-discharge plans with mothers to start their healthy transition of preterm neonate to home and to ameliorate family concerns (11).

The health interventions strategies should involve health education that is simple to conduct, utilize and cost effective. However, to date, a specific, accurate, and easy-to-follow health information regarding premature baby care is limited in Tanzania.

This study therefore, aims to introduce an educational tool in form of a brochure that can be used to train mothers of preterm infants on breast feeding, how to properly conduct kangaroo mother care (KMCI as well as the importance of hygiene practices to increase knowledge of mothers in taking care of their infants post discharge and hence save lives of preterm babies.

## Theoretical model; The Social Cognitive theory (SCT)

The Social Cognitive theory (SCT) initially called the Social Learning theory developed by Albert Bandura in the 1960s and was later modified into the Social Cognitive Theory in 1986. This theory will be applied to guide this study, it is made up of five components that affect the likelihood for an individual to change and or influence health behavior. These components include behavior capability, self-efficacy, expectations and goals, reciprocal determination, modeling and reinforcement. However for the purpose of this study only three main factors namely behavior capability, self-efficacy, and expectations and goal from this theory will be integrated.

### Self-Efficacy

This theory explains that in order to foster behavior change there must be a fostering of mothers’ belief in their ability to carry out practices successfully. Behavioral Capability: Currently mothers are lectured on the importance of breastfeeding, KMC and hygiene when taking care of their infants (9). Expectations and Goals: the SCT explains that setting realistic expectations and achievable goals enable better commitment. The brochure aims to break down these practices into manageable steps and appreciating their accomplishments during the weekly follow-up in hopes to reinforce their commitment on breastfeeding duration, frequency of KMC, and hygiene routines even post discharge.

**Figure 1:**
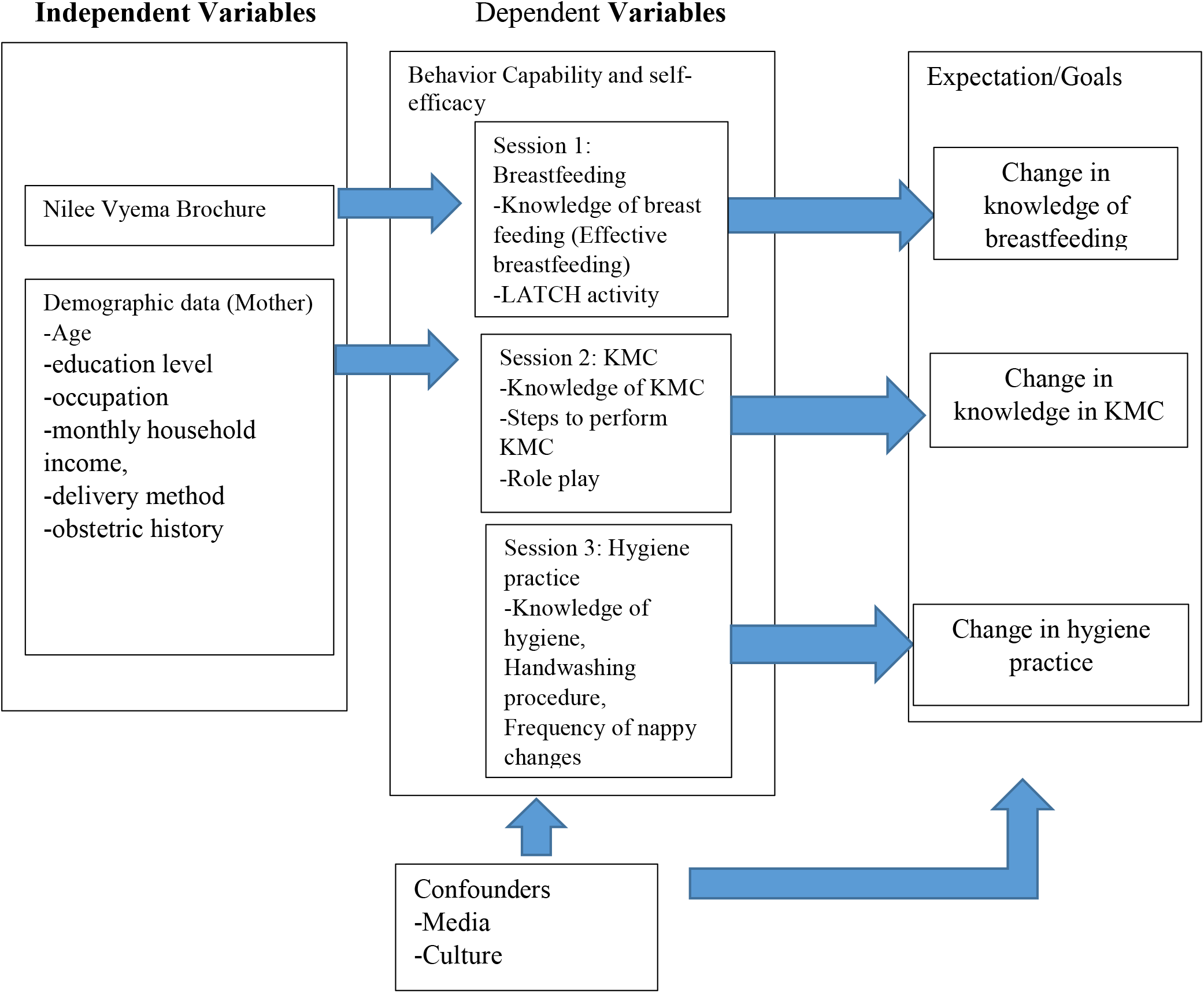
Social Cognitive Model for assessing effectiveness of Nilee Vyema Brochure on Knowledge of Breastfeeding, kangaroo mother care and hygiene practice.

## Methods

### Study Aim

The aim of this study is to assess the effectiveness of NILEE VYEMA brochure on knowledge of breastfeeding, kangaroo mother care and hygiene among mothers with preterm infants in central Tanzania.

### Study design

A two group quasi-pretest posttest experimental design will be used in this study

### Study area

This study will be conducted in central Tanzania. Central Tanzania is made up of three regions namely Dodoma, Singida and Tabora regions. The study setting will involve two regional hospitals whereby two hospitals will be randomly selected one hospital will be intervention group and one hospital will be the control group. Dodoma regional referral hospital in Dodoma provides healthcare services to a total of seven districts namely; Dodoma, Kongwa, Chamwino, Bahi, Kongwa, Mpwapwa, and Chemba. Singida regional referral hospital in Singida Region provides healthcare services to a total of five districts namely Ikungi, Singida, Manyoni, Iramba, and Mkalama. Kitete Regional Referral Hospital in Tabora provide healthcare and referral services to seven districts namely; Igunga, Kaliua, Nzega, Tabora, Urambo and Uyui. All regional hospitals in central Tanzania have a full functioning and well equipped neonatal intensive care unit (NICU).

### Study population

The study population will involve mothers with preterm infants who are admitted at the NICU.

### Sample size estimation

The sample size will be estimated using the West and Briggs formula (12) for quasi experiments using two groups. Proportion of mothers’ knowledge at pre-intervention of 77.16% (13), proportion of mothers after an intervention of 90.33% (13), standard normal deviate of 0.84 with a power of demonstrating a statistically significant difference before and after the intervention between two groups at 90%, standard normal deviation of 1.96 at 95% confidence level will be used in this study. Hence the minimum sample size will be 112 participants and will be divided into two equal groups; 56 mothers in intervention group and 56 mothers in control group.

### Sampling procedure

Simple random sampling will be used to select two regional referral hospitals from among the three regional hospitals in central Tanzania. Of the two regional referral hospital selected the first regional hospital will be control group while the second referral hospital will be intervention group. The eligible mothers found in the NICU during the time of study will be conveniently selected to participate in the study.

### Measurement variables

#### Breastfeeding knowledge

This will refer to mother’s ability to recall and mention the importance of breastfeeding and explain how to provide successful breastfeeding session to the preterm infant. The knowledge of breastfeeding will be assessed by the LATCH assessment tool (14). The LATCH assessment tool the LATCH scoring system, which is regarded as one of the most essential tools used to assess efficiency of early breastfeeding. Each letter of the acronym LATCH denotes the key component which is the area of assessment. L - how well the infant latches onto the breast. A - the amount of audible swallowing noted. T - the mother’s nipple type. C - mother’s level of comfort. H - amount of help the mother needs to hold her infant to the breast. The system assigns a numerical score, 0, 1, or 2, to five key components of breastfeeding. The mothers will be asked to respond on questions about state the essentials skills needed for a successful breastfeeding session and will score 2 points for each correct response. A total of 10 points will be scored for all correct responses whereby a cutoff point of > 8 will indicated good breastfeeding LATCH knowledge and any score below this will be considered ‘poor breastfeeding knowledge’.

#### Knowledge of KMC

This will assess the overall mother’s knowledge on KMC. The meaning of KMC, to whom it is performed on, duration, its importance, when it should be done and for how long, among other items to be assessed by questionnaire adapted from a similar study (15). The mothers will be asked to respond on a total of 10 questions where, each correct response will score one mark. All mothers who score above the cut off point of 60 will be regarged as knowledgeable while those who score below this score will be regarded as not knowledgeable. The correct responses will be compared via the pre and posttest questionnaire in the intervention group and control group.

#### Knowledge of hygiene

Hygiene knowledge will be measured by using a checklist adapted from Essential Early Newborn Care (EENC) developed by WHO in 2018 which contains a total of 8 YES/NO items in nominal scale (16). The mother will be asked to mention what she will do post discharge to ensure that her infant is protected from infection of disease as required by the EENC. Each correct item will score 2 points while for each wrong item will score 0 points. The mother can score a total of 16 points where a cutoff point of > 8 points will be used to be judge hygiene knowledge status as good. Any score below 8 will be judged as having poor knowledge.

The demographic characteristics of mothers will include age, educational level, occupation, monthly household income, delivery method, and obstetric history. The demographic characteristics of infants will include gestational age, sex, birth order, and length of hospital stay in days. These information will be obtained by face to face interview with the mothers while in the ward.

### Data collection tools and data collection methods

In this study, several methods of data collection methods will be used depending on the type of data needed to answer a particular study objective as follows:

#### Interview

A researcher-administered questionnaire will be used to obtain data from the mothers for assessing their knowledge and skills on the breastfeeding, and KMC. The data will be collected for 10 minutes.

#### Self-reported checklist

Interview will be used to ask the mother various questions regarding hygiene practice. The mother will be required to mention in detail a list of things she will do to ensure her preterm infant does not get infected with communicable diseases post discharge.

In order to avoid hospital bias, the data collector will not be one of the nurses working in the chosen facility: respondents’ confidentiality will be assured.

### Data management plan

Data obtained from participants will be stored in a secured computer with password that only the researcher’s team will be able to access, and all University of Dodoma safety protocols will be followed in order to secure participants information. The data will be cleaned and checked for completeness before being subjected to models of analysis.

### Data analysis plan

The Data will be analyzed using IBM SPSS version 25. Data will be analyzed for descriptive and inferential statistics. Frequency and percentage will be computed to describe characteristics of the study population while. Chi square will be computed to determine relationship between categorical variables at p <0.05.

### Ethical consideration and declaration

Ethical permission for this study was obtained from the University of Dodoma Research Ethics Committee (UDOM-REC) with reference number MA.84/261/70/30, Singida Regional referral hospital with a reference number MB.295/345/01/164 and Dodoma Regional Referral hospital with a reference number. A written consent will be given to the mothers of the preterm infants after explaining the purpose of the study and being told that their participation is voluntary and they can withdrawal from the study at any time they please.

### Status and timeline of the study

The study is expected to be conducted for a period of four weeks, and on the day of manuscript submission, the status of the study was in data collection plans.

### Strength and limitation

#### Strength

The strength of this study is that it will introduce an innovative method for providing health education to the mothers with preterm infants admitted in the wards at regional referral hospitals in central Tanzania. The information generated will serve as basis for describing how this change in modality will influence mother’s knowledge of caring for preterm infants’ post discharge. To the best of the authors’ knowledge, this will be the first study of its kind to be conducted to this vulnerable population in central Tanzania. Furthermore, the information may help the Ministry of Health make proper informed and innovative decisions towards preterm services and access to health information for mothers and legal guardians with preterm infants post discharge. All this aim to contribute to increase mother’s knowledge of care hence better health outcomes to the preterm infant.

#### Limitations

This study faces the following limitations; first it deals with mothers with preterm infants who will be stable during the time of study. This means that mothers with preterm infants who will not stable during the time of study, male parents and legal guardians will not be included in this study.

Thus the findings cannot be generalized to the male parent or other legal guardians of the preterm infant. The second limitation is that the study is conducted in only two regional hospitals out of three in central Tanzania therefore the findings represent the population that had preterm infants in a specific NICU at a particular time of the year and cannot be generalized to other NICUs in Tanzania; hence the findings can only be representatives of these two regions.

### Dissemination plan

Findings obtained from this study will be disseminated to the University of Dodoma, the Ministry of health, Presidents Office-Regional Administration and Local Government and will be submitted for publication in peer-reviewed journal.

### How amendments to the study, including termination, will be dealt with

If any changes are made to the study, including the study’s termination, this will be communicated to the journal auditoria, which will submit the changes or provide a reason for the change.

## Data Availability

If no pilot data are reported in the manuscript, please enter the following: All relevant data from this study will be made available upon study completion.

## Authors’ contribution

Writing – original draft, conceptualization and methodology: **Shimwe Amos Ngeze**

Writing – review, editing and supervision: **Julius Ntwenye, Richard Mongi**

## Supporting information

Knowledge, Attitude and Practice quesrionnaire (15)

Early Essential Newborn Care (EENC) (16)

LATCH Assessment tool (14)

